# Peripheral Airway Dysfunction in Symptomatic Gastroesophageal Reflux Disease: A Laboratory-Based Study Using Impulse Oscillometry

**DOI:** 10.64898/2026.08.24.26361198

**Authors:** Thilini Illangasinghe, Niranga Manjuri Devanarayana, Dilesha Wadasinghe, Manori Vijaya Kumari

## Abstract

**Introduction:** Individuals with Gastroesophageal Reflux Disease (GERD) often experience airway inflammation and bronchoconstriction as a result of reflux aspiration and/or vagally mediated reflexes. The Impulse Oscillometry System (IOS) is a sensitive, non-invasive tool that can detect subtle changes in airway resistance. While there are few studies exploring airway resistance in GERD globally, no studies have been conducted in Sri Lanka. Therefore, we aim to compare the airway resistance using IOS in medical undergraduates with and without symptomatic GERD.

**Methods:** A cross-sectional study was conducted among 811 medical undergraduates (31.1% male; mean age 22.9 years) at the Faculty of Medicine, Rajarata University of Sri Lanka. Symptomatic GERD was screened using the validated GerdQ, and a cutoff of≥8 was used to diagnose those with GERD symptoms. Of the 242 (29.8%) with GERD symptoms, 188 with chronic respiratory diseases or recent respiratory symptoms were excluded, and 50 with GERD symptoms and 50 healthy, age- and sex-matched controls were recruited. Lung function was assessed using IOS and spirometry, according to American Thoracic Society (ATS) and European Respiratory Society (ERS) guidelines.

**Results:** Prevalence of symptomatic GERD among medical undergraduates was 29.8% (242/811). The common symptoms among GERD were heartburn (89.6%, 217/242) and regurgitation (85.5%, 207/242). Oscillometry parameters including, R5-R20 Hz (15.29% vs 9.69%, p=0.002), Fres (14.95 1/s vs 13.37 1/s, p = 0.04), and AX (0.66 vs 0.48, p = 0.02) were significantly higher in students with symptomatic GERD (mean = 15.29%) than in healthy controls (mean = 9.69%; p = 0.002). However, spirometry parameters including FEV1, FVC, and PERF did not differ between the GERD-positive and control groups.

**Conclusion:** Individuals with symptomatic GERD demonstrated a higher peripheral airway resistance compared to controls, whereas no significant difference was observed in upper airway resistance. This could be due to the gastric acid stimulation of vagal nerve terminations in the lower part of the esophageal wall, leading to increased resistance in the peripheral airways through vagally mediated bronchoconstriction.

**What is already known on this topic:**

- GERD is associated with extra-esophageal conditions, including chronic cough, asthma and laryngitis.
- Pathological acid reflux can induce respiratory symptoms.

**What this study adds:**

- Individuals with symptomatic GERD demonstrated a higher peripheral airway resistance compared to those without GERD, but no difference in upper airway resistance.
- This may be attributable to vagally mediated reflex pathways, in which esophageal stimulation triggers an esophagobronchial reflex, leading to increased parasympathetic activity and subsequent bronchoconstriction.
- The novelty of incorporating impulse oscillometry provides valuable baseline data for the Sri Lankan population and contributes to the limited global literature on small airway dysfunction in individuals with GERD.
- By combining impulse oscillometry with spirometry, this study detected changes in peripheral airway resistance that may not be identified by conventional spirometry alone.

**How this study might affect research, practice or policy:**

- These findings highlight the potential value of impulse oscillometry for detecting subtle peripheral airway abnormalities that may not be apparent from conventional pulmonary function assessment.
- Further longitudinal and mechanistic studies are warranted to clarify the underlying pathways linking GERD with peripheral airway dysfunction
- In clinical practice, assessment of peripheral airway function may be considered in patients with symptomatic GERD who have respiratory complaints
- Future longitudinal studies should determine whether GERD treatment can modify peripheral airway resistance and whether such abnormalities have clinical or prognostic significance.

## INTRODUCTION

Gastroesophageal reflux disease (GERD) is a common gastrointestinal disorder characterized by the reflux of gastric contents into the esophagus (1). It is one of the most frequently encountered gastrointestinal disorders worldwide and represents a considerable health burden (2). A systematic review reported that the global prevalence of GERD was 13% (2) while a countrywide study reported that 25.3% of Sri Lankans had probable GERD (3). The pathogenesis of GERD is multifactorial and involves several abnormalities in the normal reflux mechanisms, including an increase in the duration of lower esophageal sphincter relaxation, a decrease in the pressure of the lower esophageal sphincter, esophageal acid clearance interruption, hiatal hernia, slow gastric passage, and increased gastric acid secretion (4). These physiological disturbances can result in persistent reflux symptoms and may contribute to the development of both gastrointestinal and extra-esophageal manifestations. GERD is associated with various extra-esophageal conditions, including chronic cough (5), asthma (6), recurrent pneumonia, sinusitis (7), laryngitis (8), subglottic stenosis, laryngeal cancer, non-cardiac chest pain (9,10) and esophageal stricture (11). The relationship between GERD and respiratory disorders has been increasingly recognized in recent years. A systematic review reported that there was a significant association between asthma and GERD (12). Similarly, a study conducted in Nigeria reported that 17.4% of medical undergraduates have both asthma and GERD (13).

Pathological acid reflux can trigger asthma by two complementary mechanisms. Either refluxed gastric contents may reach the larynx and lower airways through microaspiration, where direct exposure of the airway epithelium to acidic or other gastric contents can induce airway inflammation and bronchoconstriction (14), or acid exposure of the distal esophageal mucosa may stimulate esophageal sensory afferent nerves, particularly vagal nerve endings, triggering an esophagobronchial reflex that increases parasympathetic activity and promotes bronchoconstriction (15). These mechanisms may contribute to airway hyperresponsiveness and the worsening of respiratory symptoms in individuals with GERD. Although the association between GERD and respiratory disorders has been increasingly recognized, the mechanisms underlying the effects of GERD on airway function remain incompletely understood. Interestingly, a laboratory study conducted in Sri Lanka among asthmatic patients using conventional spirometry reported that distal esophageal acid exposure can increase parasympathetic activity and cause bronchoconstriction in patients with asthma, and strengthens the hypothesis that the gastroesophageal reflex may trigger asthma-like symptoms through a vagally mediated esophagobronchial reflex (16).

GERD may influence respiratory function even in the absence of prominent respiratory symptoms. However, conventional pulmonary function tests such as spirometry primarily assess airflow and may have limited sensitivity for detecting subtle changes in the peripheral airways (17). In contrast, impulse oscillometry (IOS) detects peripheral airway abnormalities that are not readily apparent on conventional spirometry (18). Therefore, IOS may be a valuable complementary technique for investigating subtle alterations in airway function associated with GERD. Therefore, this study evaluated lung function in individuals with GERD using impulse oscillometry, which is more sensitive than spirometry in assessing direct airway resistance and subtle airway changes.

## METHODS

### Study Design and Setting

This study was conducted among medical undergraduates in the Faculty of Medicine and Allied Sciences at Rajarata University of Sri Lanka. The study consisted of two phases. Phase I was a questionnaire-based cross-sectional study to identify students with symptomatic GERD. Students who were identified as having symptomatic GERD in phase I were invited to a laboratory-based study to assess the lung function, which was phase II of this study.

### Sample size calculation

In phase I, the sample size was calculated to determine the prevalence of GERD among medical undergraduates using the formula n=Z^2^P(1−P)/d^2^, with an absolute precision of 5% and a standard normal deviation of 1.96 for a 95% confidence level. P is the expected prevalence, which was used as 25.3%, since a countrywide study reported that 25.3% of Sri Lankans had probable GERD (3). The calculated minimum required sample size was 290, and considering non-responders (60%), the final sample size was determined as 484.

In phase II, the laboratory-based study, there were two groups: symptomatic GERD-positive and symptomatic GERD-negative. The sample size was calculated using Win Pepi statistical software (19), assuming a two-sided significance level of 0.05, 80% statistical power, and an expected standardized mean difference of 1.0. The calculated minimum sample size was 18 participants in each group (symptomatic GERD-positive and symptomatic GERD-negative), giving a total minimum sample size of 36 participants.

### Ethical Approval and Informed Consent

Ethical approval was obtained from the Ethics Review Committee, Faculty of Medicine and Allied Sciences, Rajarata University of Sri Lanka (ERC/2024/60). Informed written consent was obtained from the participants prior to the participation. Undergraduates were allowed to withdraw voluntarily from the study at any point without providing a reason, and this did not affect their academic work in any manner. Participation was voluntary and anonymous, with no academic incentives. Administrative clearance was obtained from the Dean of the Faculty of Medicine and Allied Sciences, Rajarata University of Sri Lanka. A separate identification number was assigned to each undergraduate, and the subjects’ socio-demographic details were kept strictly confidential. Data was stored as a computerized package with a password protected document, which is accessible only to investigators.

### Data Collection of cross-sectional study (Phase I)

Phase I was conducted using a self-administered questionnaire, distributed in online platform. Before distribution, the questionnaire was pretested with 30 medical undergraduates to assess feasibility and identify unclear items. All questionnaires were administered in English language as medical degree programme is conducted in English language, and undergraduates are familiar with medical term. Additional clarification provided during the data collection process if any students request it. A pretest was conducted with the participation of 30 medical undergraduates to assess the feasibility of filling the questionnaire. and find any problems faced due to unclear words or unclear meanings. Any errors were revised, re-drafted & finalized before administering the final questionnaires.

#### Diagnostic tool to identify symptomatic GERD

The validated Gastroesophageal Reflux Disease Questionnaire (GerdQ) (20) was used to identify individuals with GERD. GerdQ comprises 6 items and assesses symptoms of GERD over the previous 7 days. A four-point Likert scale (0–3) is used to score the frequency of four positive predictors of GERD, and a reverse Likert scale (3–0) is used for two negative predictors of GERD (epigastric pain and nausea), yielding a total GERD score range of 0–18. The diagnosis of probable GERD was made if the composite GERD score cut-off was 8 or greater.

#### Asthma screening tool

A validated symptoms-based screening questionnaire (Asthma Screening Questionnaire-ASQ) (21) was used to screen the students with probable asthma. This is a simplified 6-item questionnaire, which includes cough, chest tightness, wheezing, and shortness of breath. The total ASQ score is calculated by summing all positive responses, yielding a patient score ranging from 0 to 20. An ASQ cutoff score ≥4 was used to screen students with asthma.

#### Data collection of laboratory-based study (Phase II)

Students who met the criteria for symptomatic GERD during the survey were subsequently invited to participate in the laboratory-based study. Fifty-six students, identified as having symptomatic GERD (symptomatic GERD positive) during phase I, were recruited for a laboratory-based study, provided they did not have any chronic respiratory or gastrointestinal disorders other than GERD (Figure 1). Age and sex-matched healthy students (n=75) without GERD or respiratory symptoms, and with no history of chronic respiratory or gastrointestinal disorders, were recruited as the control group (GERD negative). Both groups underwent impulse oscillometry and spirometry in accordance with American Thoracic Society (ATS) and European Respiratory Society (ERS) guidelines (22,23). The lung function reports were evaluated by both an expert respiratory physiologist and a consultant respiratory physician. The reports confirmed as accurate and reliable by both experts were included in the final analysis. (GERD case; n=50, controls; n=50)

**Figure 1.**
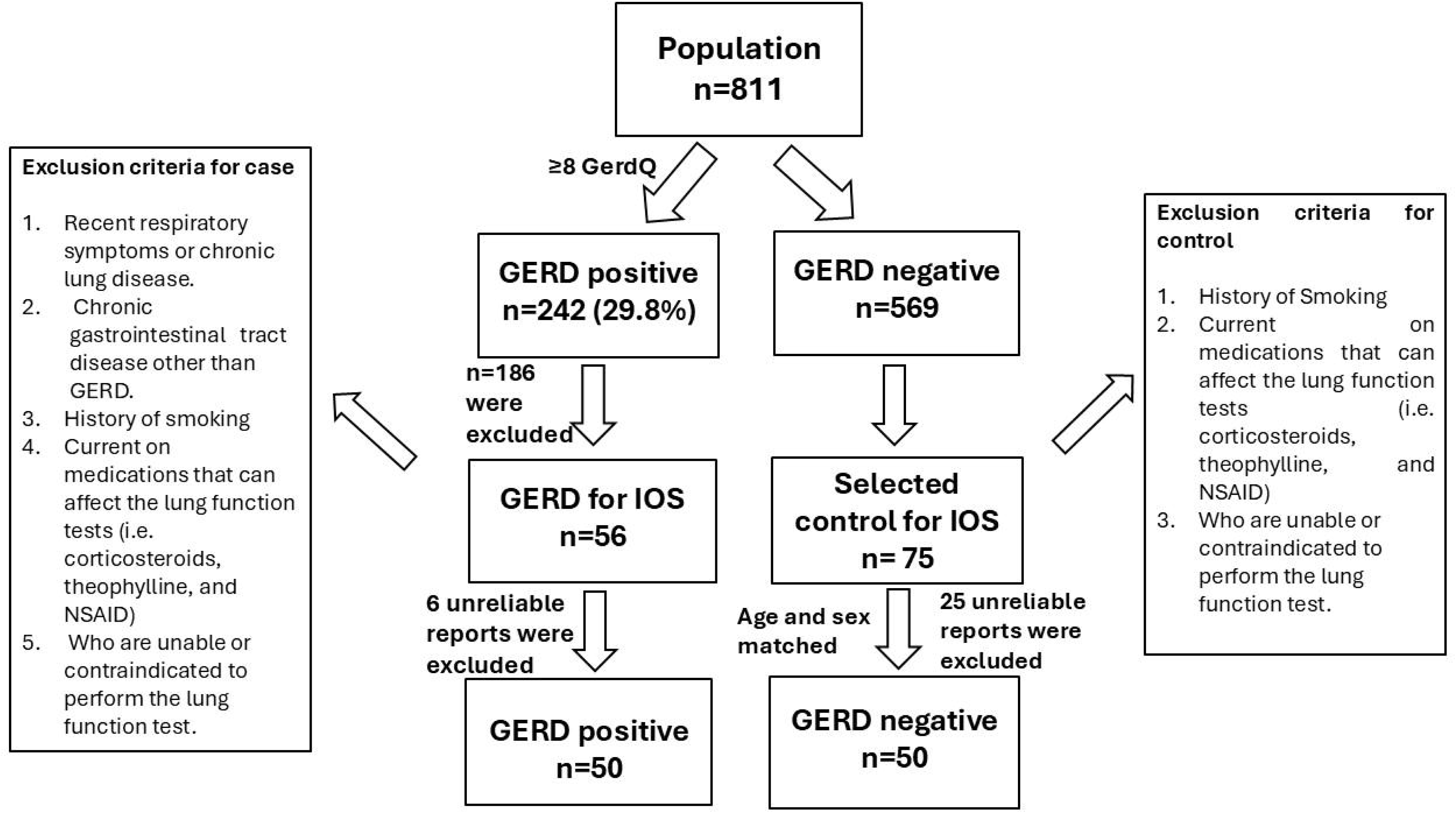
Study flow diagram showing participant selection and recruitment for pulmonary function test by impulse oscillometry and spirometry

### Assessment of lung function parameters

Impulse oscillometry and spirometry were performed following American Thoracic Society and European Respiratory Society guidelines (22). A calibration check was performed prior to each test using a 3L calibration syringe. Disposable bacterial filters were used during each measurement to prevent cross-infection. The manoeuvre was first demonstrated to the medical undergraduate by the investigator. The subjects were then asked to sit during the procedure. A nose clip is applied to prevent breathing through the nose.

During spirometry, the subjects were asked to take a deep breath and then hold the mouthpiece between their lips to create a good seal. They were instructed to expire forcefully and maximally until they feel that no breath is left and then inspire rapidly to maximum capacity. The subjects were allowed to rest between manoeuvres as repeated maximal efforts could trigger bronchospasm. Each manoeuvre was monitored using flow-volume loops and manual observation to ensure that the effort was maximal, smooth, and artefact-free. Three reproducible replicates were considered acceptable, and the highest record was obtained from the acceptable replicates. The following parameters were recorded after each attempt: Forced Vital Capacity (FVC), Forced Expiratory Volume in the first second (FEV1), Forced Expiratory Flow between 25-75% (FEF 25%-75%), Peak Expiratory Flow Rate (PEFR).

Impulse oscillometry (IOS) is a non-invasive test (24) based on the forced oscillation principle, in which sound waves of varying frequencies (typically 4-50 Hz) are superimposed on normal breathing to assess airway function (22). It reports resistance (Rrs) and reactance (Xrs) at different frequencies (25). During impulse oscillometry, the subject was instructed to breathe quietly while maintaining a tight seal around the mouthpiece. During breathing, the cheeks and floor of the mouth were supported firmly using both hands. During the test, the subject was instructed to avoid swallowing, relax the tongue, and keep it below the mouthpiece. Three replicate measurements were obtained and considered acceptable if they met specified quality criteria, including visual inspection, and a within-session coefficient of variability (CoV) of less than 10% (22). The following parameters were recorded after each attempt: total airway resistance (Rrs5), central airway resistance (Rrs20) and peripheral airway resistance (Rrs5-Rrs20), total reactance at all frequencies between 5 Hz and Fres (AX) and resonant frequency (Fres) (26).

### Statistical analysis

Data was analyzed using SPSS software (version 26). Binary logistic regression was used to assess the association between GERD and asthma. Chi-square test was used to identify the associated gastrointestinal symptoms and asthma. The independent-sample t-test was used to compare lung function parameters between undergraduates with and without symptomatic GERD. The Spearman correlation coefficient was used to assess the correlation between lung function parameters and GERD score.

## RESULTS

### Sample Characteristics

A total of 959 questionnaires were distributed among medical undergraduates, and 816 (85%) completed questionnaires were returned. Of them, 811 (99.3%) were properly completed and were included in the final analysis. The final sample included 559 (68.9) females and 252 (31.1%) males, with a mean age of 22.93 years (SD 2.09 years) (Table 1).

**Table 1:**
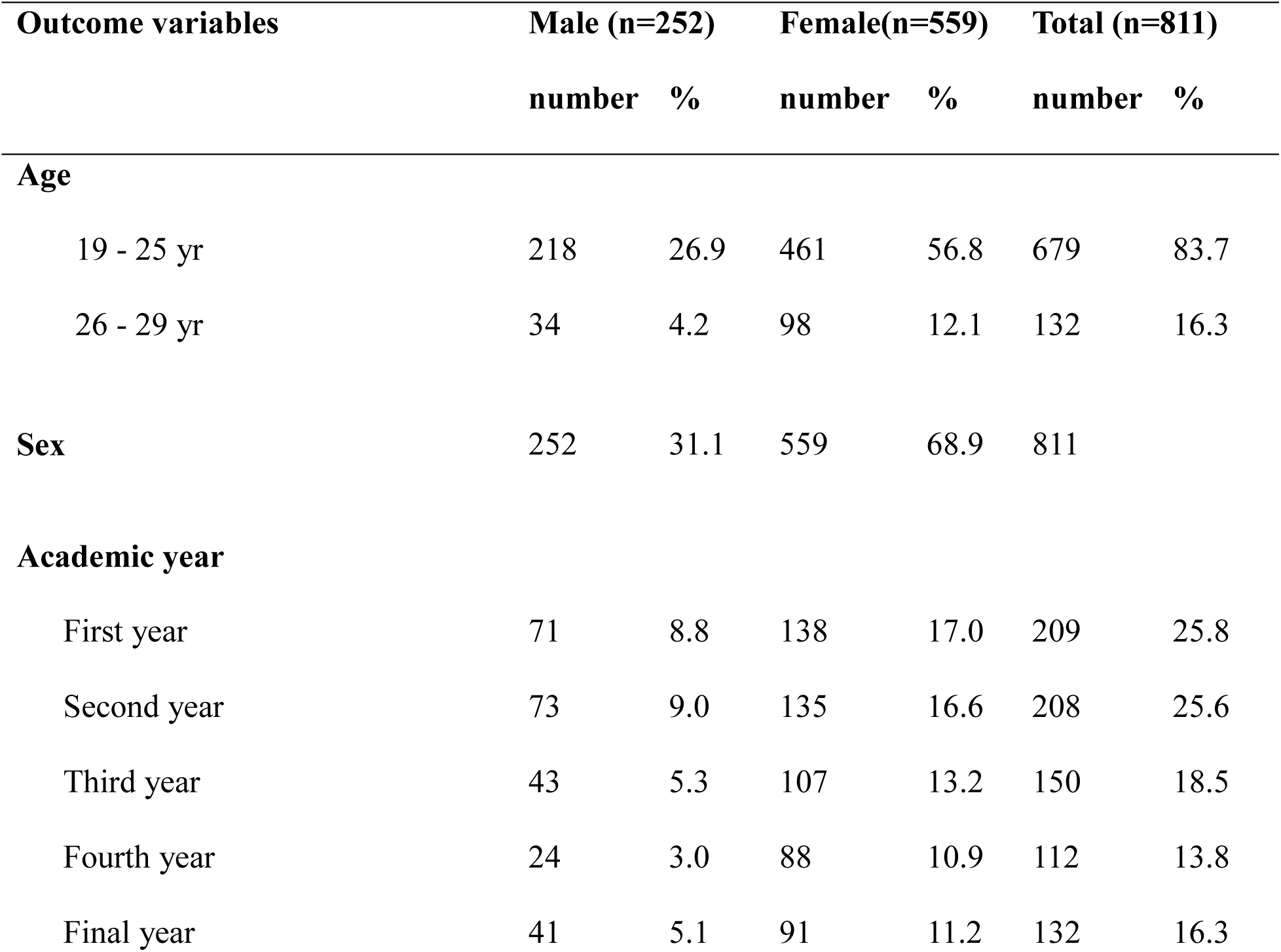
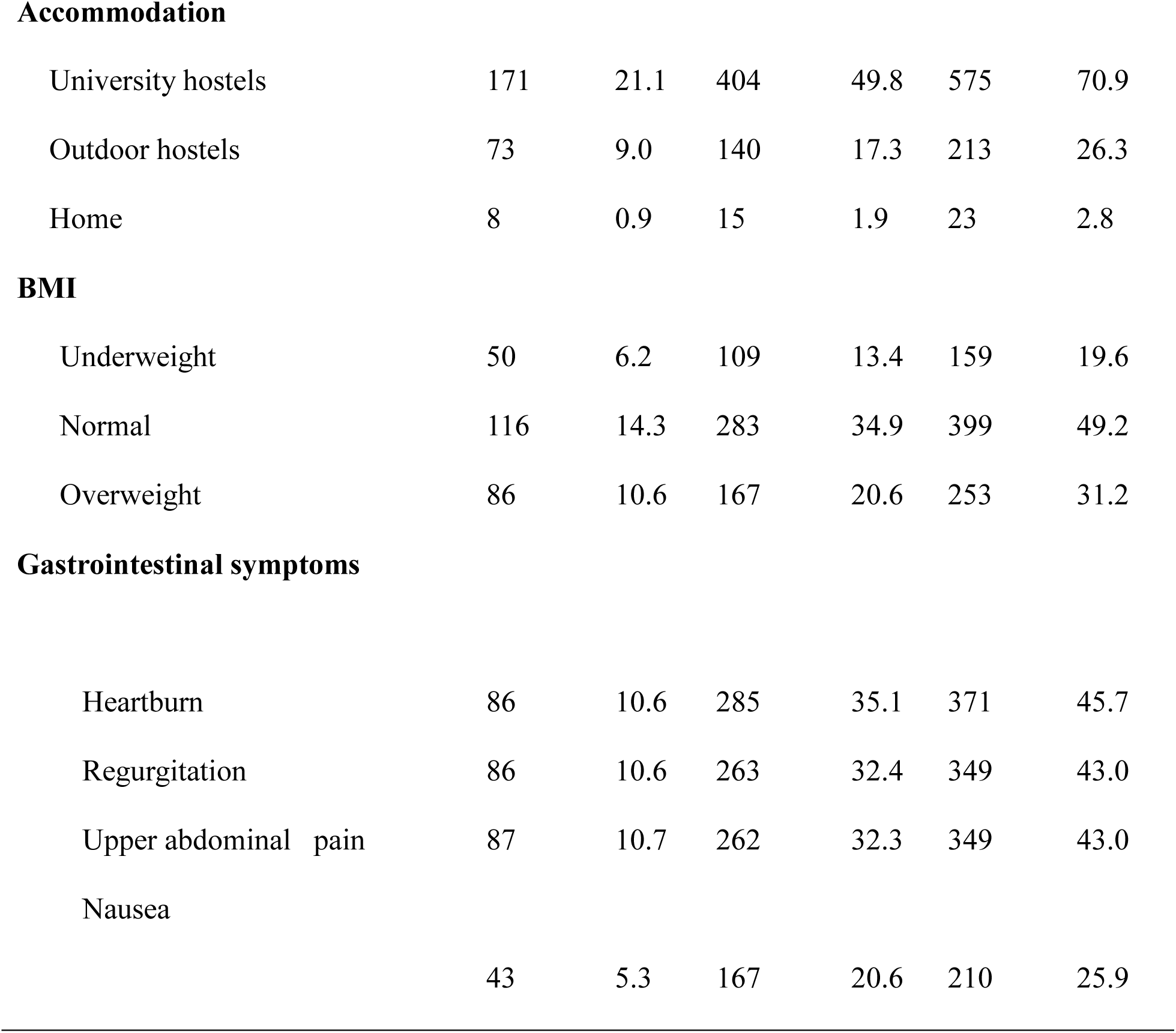
Sociodemographic distribution of the study sample.

| Outcome variables | Male (n=252) |  | Female(n=559) |  | Total (n=811) |  |
| --- | --- | --- | --- | --- | --- | --- |
|  | number | % | number | % | number | % |
| **Age** |  |  |  |  |  |  |
| 19 - 25 yr | 218 | 26.9 | 461 | 56.8 | 679 | 83.7 |
| 26 - 29 yr | 34 | 4.2 | 98 | 12.1 | 132 | 16.3 |
| **Sex** | 252 | 31.1 | 559 | 68.9 | 811 |  |
| **Academic year** |  |  |  |  |  |  |
| First year | 71 | 8.8 | 138 | 17.0 | 209 | 25.8 |
| Second year | 73 | 9.0 | 135 | 16.6 | 208 | 25.6 |
| Third year | 43 | 5.3 | 107 | 13.2 | 150 | 18.5 |
| Fourth year | 24 | 3.0 | 88 | 10.9 | 112 | 13.8 |
| Final year | 41 | 5.1 | 91 | 11.2 | 132 | 16.3 |
| **Accommodation** |  |  |  |  |  |  |
| University hostels | 171 | 21.1 | 404 | 49.8 | 575 | 70.9 |
| Outdoor hostels | 73 | 9.0 | 140 | 17.3 | 213 | 26.3 |
| Home | 8 | 0.9 | 15 | 1.9 | 23 | 2.8 |
| **BMI** |  |  |  |  |  |  |
| Underweight | 50 | 6.2 | 109 | 13.4 | 159 | 19.6 |
| Normal | 116 | 14.3 | 283 | 34.9 | 399 | 49.2 |
| Overweight | 86 | 10.6 | 167 | 20.6 | 253 | 31.2 |
| **Gastrointestinal symptoms** |  |  |  |  |  |  |
| Heartburn | 86 | 10.6 | 285 | 35.1 | 371 | 45.7 |
| Regurgitation | 86 | 10.6 | 263 | 32.4 | 349 | 43.0 |
| Upper abdominal pain | 87 | 10.7 | 262 | 32.3 | 349 | 43.0 |
| Nausea | 43 | 5.3 | 167 | 20.6 | 210 | 25.9 |

### Prevalence and symptomatology of gastroesophageal reflux disease

The prevalence of symptomatic GERD was 29.8% (242/811). The most common symptoms among medical students with GERD were heartburn (89.6%, 217/242), regurgitation (85.5%, 207/242), epigastric pain (63.6%, 154/242), and nausea (35.9%, 87/242). Nearly half of those with GERD symptoms had sleep disturbance (47.5%, 115/242) and had used self-medication (50.8%, 123/242) at least one day per week to relieve their symptoms.

### Gastrointestinal symptoms among individuals with asthma

The prevalence of asthma was 18.5% (150/811) among medical undergraduates. Sixty-one undergraduates (7.5%, 61/811) had both asthma and GERD. There was an independent association between asthma and GERD in medical undergraduates (adjusted OR 1.79, 95% CI 1.24-2.60, *p*=0.002, logistic regression adjusted for age and sex). Gastrointestinal symptoms, including heartburn (adjusted OR 1.65, 95% CI 1.14-2.39, *p=*0.007), regurgitation (adjusted OR 1.91, 95% CI 1.33-2.74, *p*=0.0005), upper abdominal pain (adjusted OR 1.90, 95% CI 1.32-2.73, *p*=0.0005), nausea (adjusted OR 2.12, 95% CI 1.45-3.12, *p*=0.0005) were more prevalent among those with asthma compared to non-asthmatics (Figure 2).

**Figure 2.**
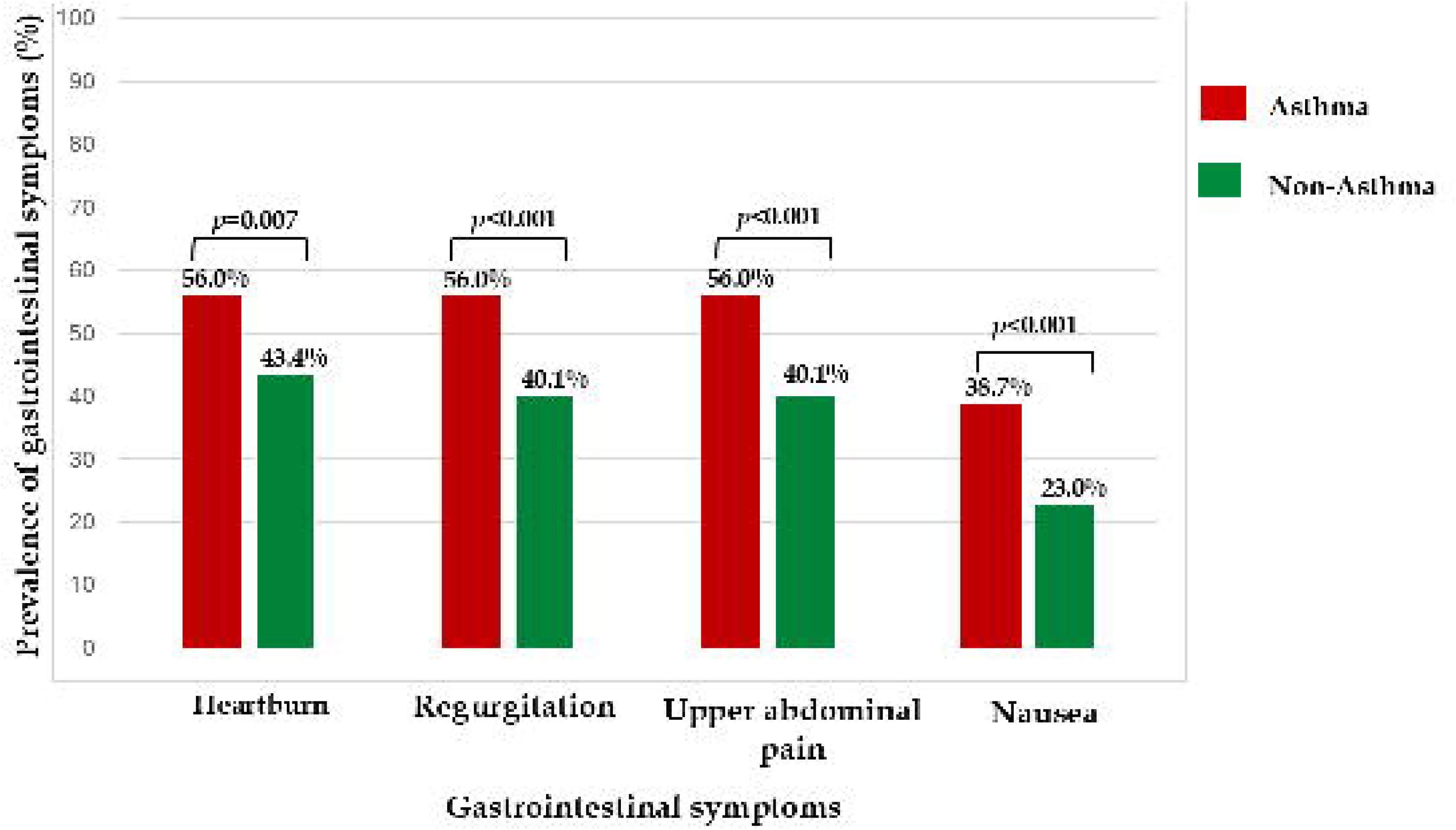
Prevalence of gastrointestinal symptoms among medical undergraduates with asthma

### Comparison of lung function parameters between individuals with and without GERD symptoms

Spirometry parameters did not differ between GERD-positive and GERD-negative groups. However, IOS parameters measuring peripheral airway resistance, including Delta R5Hz-R20Hz, Fres, and Axe, were significantly increased among individuals with symptomatic GERD compared to GERD-negative individuals (Table 2). Further, the GERD score showed a significant positive correlation with Delta R5Hz-R20Hz, which measures peripheral airway resistance (r=0.23, *p*=0.018) (Figure 3).

**Figure 3.**
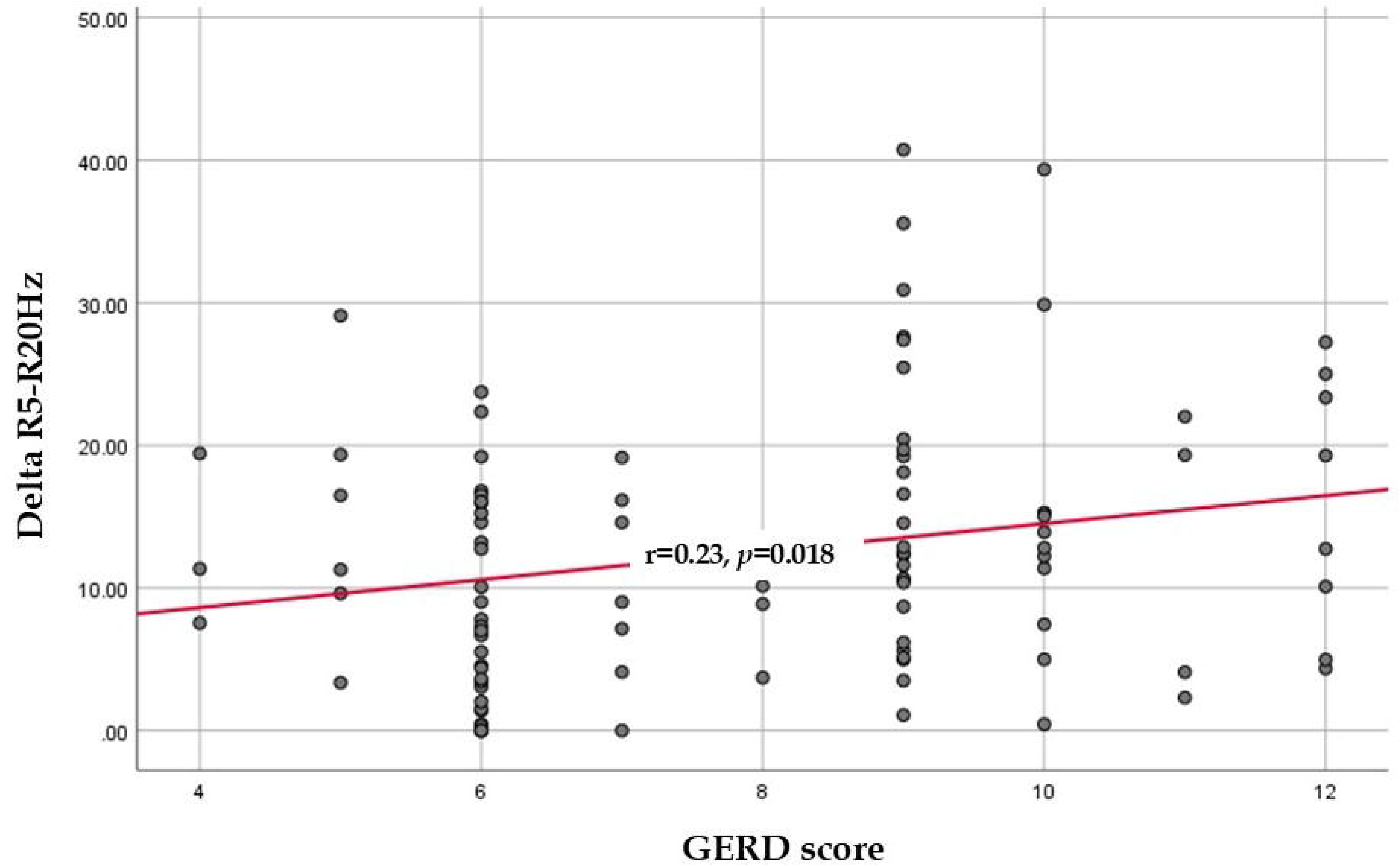
Correlation between GERD symptom score and peripheral airway resistance (Delta R5Hz-R20H), Delta R5Hz-R20Hz: peripheral airway resistance; GERD: Gastroesophageal reflux disease

**Table 2.** Comparison of lung function parameters between medical undergraduates with and without GERD symptoms.

| Parameter | GERD symptoms<br>positive (n=50)<br>mean (SD) | GERD symptoms<br>negative (n=50)<br>mean (SD) | P value |
| --- | --- | --- | --- |
| **Spirometry parameters** |  |  |  |
| FVC (L) | 2.94 (0.64) | 2.89 (0.81) | 0.7 |
| FEV<sub>1</sub> (L) | 2.61 (0.54) | 2.61 (0.68) | 0.9 |
| FEV<sub>1</sub>/FVC ratio (%) | 88.27 (6.07) | 90.15 (6.21) | 0.1 |
| PEF (L/s) | 5.15 (1.60) | 5.56 (2.13) | 0.2 |
| FEF<sub>50%</sub> (L/s) | 1.96 (0.46) | 1.98 (0.53) | 0.8 |
| **Impulse oscillometry parameters** |  |  |  |
| R5Hz (kPa·s/L) | 0.39 (0.11) | 0.37 (0.07) | 0.1 |
| R20Hz (kPa·s/L) | 0.33 (0.08) | 0.33 (0.06) | 0.9 |
| R10Hz (kPa·s/L) | 0.34 (0.1) | 0.32 (0.06) | 0.2 |
| Delta R5Hz-R20Hz<br>(kPa·s/L) | 15.29 (9.89) | 9.69 (7.22) | 0.002 |
| X5Hz (kPa/L) | -0.14 (0.04) | -0.13 (0.04) | 0.3 |
| X10Hz (kPa/L) | -0.04 (0.04) | -0.03 (0.02) | 0.01 |
| X20Hz (kPa/L) | 0.04 (0.03) | 0.05 (0.03) | 0.1 |
| Fres (Hz) | 14.95 (4.32) | 13.37 (3.35) | 0.043 |
| AX (kPa/L) | 0.66 (0.51) | 0.48 (0.24) | 0.026 |

FVC-Forced vital capacity, FEV<sub>1</sub>-forced Expiratory Volume in 1 Second, FEV<sub>1</sub>/FVC-Forced Expiratory Volume in 1 Second to Forced Vital Capacity Ratio, PEF-Peak Expiratory Flow, FEF<sub>50%</sub> -Forced Expiratory Flow at 50%, R5Hz – Resistance at 5 Hz, R20Hz-Resistance at 20 Hz, R10Hz-Resistance at 10 Hz, X5Hz-Reactance at 5 Hz, X10Hz-Reactance at 10 Hz, X20Hz – Reactance at 20 Hz, Fres – Resonant Frequency, Ax – Area of Reactance

## DISCUSSION

To our knowledge, this is the first study in Sri Lanka to assess lung function parameters using impulse oscillometry in individuals with GERD symptoms. In this study, the prevalence of symptomatic GERD among medical undergraduates was 29.8%, with heartburn and regurgitation being the most common symptoms. Coexistence of asthma and GERD was observed in 7.5%, showing a significant association between the two conditions. In the laboratory-based study, spirometry parameters did not differ between GERD-positive and GERD-negative individuals. However, impulse oscillometry revealed significantly higher peripheral airway resistance (R5– R20, Fres, Ax) in those with GERD symptoms, and also GERD symptom severity positively correlated with R5–R20, indicating increased peripheral airway involvement.

This observed prevalence of GERD symptoms (29.8%) in medical undergraduates at the Rajarata University of Sri Lanka is slightly higher than the islandwide prevalence reported in a previous Sri Lankan study (25.3%) (3). The prevalence of heartburn (55%) and regurgitation (54.2%) was observed in our medical undergraduates with GERD symptoms. Our results were comparable to a study conducted in Egypt, which identified heartburn (28.4%) and regurgitation (30%) as the most prevalent symptoms among medical undergraduates. In contrast, a study conducted among medical undergraduates in Saudi Arabia reported prevalence of symptomatic GERD as14.9%, which is notably lower than the prevalence observed in our study. Moreover, this study reported that epigastric pain (1.6%) and regurgitation (1.8%) were the least frequent symptoms (27). The differences in the prevalence of symptomatic GERD and individual symptoms across studies may be explained by variations in study populations, diagnostic criteria, symptom assessment methods, and underlying lifestyle and psychosocial factors (28). Although all studies involved medical undergraduates, differences in age distribution, sex composition, academic workload, perceived stress, dietary habits, meal patterns, sleep duration, and physical activity may have contributed to the observed variation (29,30).

In the present study, the coexistence of asthma and GERD was observed in 7.5% of students, showing a significant association between the two conditions. A previous study conducted in Sri Lankan teenagers has reported a significant association between GERD and asthma (6). A study conducted among university students in Nigeria reported 17.4% of them had both asthma and GERD (13). Additionally, studies have shown that treatment of GERD is effective for managing individuals with comorbid asthma (31). Since the foregut and respiratory tract originate from a common embryonic origin (32), an association between gastroesophageal disorders and lung disease is possible. Pathological acid reflux can trigger asthma by two complementary mechanisms. Either refluxed gastric contents may reach the larynx and lower airways through microaspiration, where direct exposure of the airway epithelium to acidic or other gastric contents can induce airway inflammation and bronchoconstriction (14,33), or acid exposure of the distal esophageal mucosa may stimulate esophageal sensory afferent nerves, particularly vagal nerve endings, triggering an esophagobronchial reflex that increases parasympathetic activity and promotes bronchoconstriction (15). These mechanisms may contribute to airway hyperresponsiveness and the worsening of respiratory symptoms in individuals with GERD (34).

In our study, we assessed the lung function parameters in individuals with GERD with no respiratory symptoms using spirometry and impulse oscillometry,. In a laboratory-based comparison, spirometry parameters with respect to FVC, FEV1, FEV1/FVC ratio, PEF, and FEF_50%_ did not differ between GERD-positive and GERD-negative individuals. Similarly, a study conducted in Egypt among individuals with GERD reported no significant abnormalities in spirometry parameters with severity of GERD (35). In contrast, a study conducted in Iran among individuals with GERD with no respiratory symptoms and healthy controls reported a significant difference in FEV1, FVC using a body plethysmograph between the two groups (34). Moreover, a study conducted in Iraq reported a highly significant reduction in FEV1, PEF, and FMF, revealing an obstructive airway pattern in individuals with GERD associated cough and/or asthma compared with healthy controls (36). These inconsistent findings may be attributed to differences in study populations, particularly the presence or absence of respiratory symptoms, asthma, or GERD-related cough s, which may predispose individuals to detectable airway obstruction. Additionally, variations in GERD severity, diagnostic criteria, and lung function assessment techniques, such as spirometry versus, may also contribute to the observed discrepancies. Moreover, spirometry may be less sensitive in detecting subtel airway changes in symptomatic individuals with GERD. Therefore, in our study impulse oscillometry was used to further evaluate and detect subtle airway resistance changes that may not be identified by conventional spirometry.

In our study, impulse oscillometry revealed significantly higher peripheral airway resistance (as indicated by ΔR5–R20, Fres, Ax) in individuals with symptomatic GERD compared to control,, even in the absence of prominent respiratory symptoms. This finding was further strengthened by showing the positive correlation between GERD severity with R5–R20Hz, indicating increased peripheral airway involvement. Similar to our study findings, a study conducted in Iran reported that total airway resistance (R5Hz) was significantly higher among individuals with GERD than healthy controls (34). In contrast, another study investigated the respiratory function of GERD patients before and after omeprazole treatment. Initial assessments revealed elevated central airway resistance (R20Hz) prior to omeprazole intervention. Following treatment, improvements were observed in the R5Hz, R20Hz, and X5Hz values, suggesting that individuals with GERD may exhibit airway dysfunction, even in the absence of overt respiratory symptoms (37). One possible explanation is that increased peripheral airway resistance in individuals with symptomatic GERD is brought by a vagally mediated esophagobronchial reflex. Acid exposure in the distal esophagus can stimulate esophageal sensory afferent pathways (38), which transmit signals through the vagus nerve to the brainstem and subsequently increase parasympathetic activity to the airways, resulting in bronchial smooth-muscle constriction and increased airway resistance (39). Experimental studies have demonstrated that intraesophageal acid exposure can reduce expiratory airflow and increase airway resistance even in the absence of evidence of microaspiration, supporting the involvement of a vagally mediated reflex (40). Therefore, repeated exposure of the esophageal mucosa to gastric acid in GERD may induce reflex bronchoconstriction and contribute to increased peripheral airway resistance.

This study has several important strengths that enhance the validity and applicability of the findings. To the best of our knowledge, this is the first study conducted in Sri Lanka to assess respiratory function using impulse oscillometry in individuals with symptomatic GERD, particularly among medical undergraduates. The novelty of incorporating impulse oscillometry provides valuable baseline data for the Sri Lankan population and contributes to the limited global literature on small airway dysfunction in individuals with GERD who lack prominent respiratory symptoms. The findings add new evidence to understanding the extra-esophageal manifestations of GERD and their potential subclinical respiratory effects. Our study used both spirometry and impulse oscillometry to assess respiratory function. Spirometry is considered the gold-standard method for evaluating airflow limitation; however, it is relatively insensitive in detecting early or subtle changes in small-airway function. By combining spirometry with impulse oscillometry, this study detected changes in peripheral airway resistance that may not be identified by conventional spirometry alone. This dual-modality assessment improves the sensitivity of respiratory function evaluation and strengthens the hypothesis that individuals with symptomatic GERD may have subclinical peripheral airway involvement despite normal spirometric findings.

Our study evaluates respiratory function in individuals with GERD who do not have prominent respiratory symptoms. Many previous studies have included individuals with asthma, chronic cough, or other respiratory complaints, potentially confounding the association between GERD and respiratory function abnormalities (41–43). By focusing on a relatively asymptomatic population, this study identified subtle airway changes that may represent early or subclinical respiratory involvement. This strengthens the argument that GERD itself may contribute to airway dysfunction, independent of respiratory symptoms. Furthermore, the comparison between GERD-positive and GERD-negative individuals strengthens the study’s analytical design. This comparative approach allows identification of differences in airway resistance and improves the interpretability of findings. The inclusion of multiple IOS parameters and correlation with GERD severity further supports the robustness of the analysis.

Despite its strengths, this study has several limitations. Our study population consisted only of medical undergraduates, which may limit the generalizability of the findings to the broader population. Medical undergraduates may have unique lifestyle factors such as irregular sleep patterns, academic stress, dietary habits, and caffeine consumption, which may influence both GERD symptoms and respiratory function. Therefore, the generalizability of findings will be challenging. Notably, the diagnosis of symptomatic GERD in this study was made on a symptom-based-questionnaire rather than objective diagnostic tools such as endoscopy, pH monitoring, or impedance monitoring. According to the American College of Gastroenterology (ACG) guidelines (44), if the patient presents with classic symptoms of GERD, such as heartburn and regurgitation, endoscopy is not initially recommended to confirm GERD. After an 8-week period of Proton Pump Inhibitor (PPI) therapy, if the patient is only resistant to treatment, there will be an indication for endoscopy and PH manometry later on for the diagnosis of GERD (44). As our study did not provide empirical treatment for this population, there was no indication to perform invasive investigations such as endoscopy or pH manometry. Therefore, we assessed symptomatic GERD in this population using a validated Gerd-Q questionnaire. The study also did not include longitudinal follow-up after GERD treatment. Assessing respiratory function before and after anti-reflux therapy would help determine whether peripheral airway resistance improves. Such findings would strengthen the causal relationship between GERD and airway dysfunction. Notably, although impulse oscillometry is sensitive in detecting small airway changes, it is not as widely standardized as spirometry. Variability in IOS measurements and reference values may affect interpretation. The absence of locally derived reference values for impulse oscillometry in the Sri Lankan population may also limit the interpretation of absolute values. Establishing local reference values for impulse oscillometry in South Asian populations would also improve the accuracy of interpretation. Overall, future research integrating clinical assessment will be essential to fully elucidate the complex relationship between GERD and respiratory function.

## CONCLUSION

Individuals with symptomatic GERD demonstrated a higher peripheral airway resistance compared to healthy controls, whereas no significant difference was observed in upper airway resistance. This could be due to the gastric acid stimulation of vagal nerve terminations in the lower part of the esophageal wall, leading to increased resistance in the peripheral airways through vagally mediated bronchoconstriction.

## Acknowledgment

The authors would like to express their sincere gratitude to Professor Lakmali Amarasiri for her expertise in interpreting impulse oscillometry reports. Her guidance and expertise greatly contributed to the accurate interpretation of the respiratory function findings. The authors also wish to express their sincere appreciation to the Head, Department of Physiology, Faculty of Medicine and Allied Sciences, Rajarata University of Sri Lanka, for facilitating access to the Lung Function Laboratory. We also sincerely thank all the students who participated in the study.

## Author contributions

Thilini Illangasinghe contributed to designing the study, data collection, data curation, data visualization, data analysis, data interpretation, investigation, methodology, fund acquisition, and writing of the original draft. Niranga Manjuri Devanarayana contributed to designing the study, methodology, project administration, and critically reviewed the final manuscript for important intellectual content. Dilesha Wadasinghe contributed to methodology and critically reviewed the final manuscript for important intellectual content. Manori Vijaya Kumari contributed to conceptualization, designing the study, data curation, data visualization, data analysis, data interpretation, investigation, methodology, project administration, fund acquisition, and critically reviewed the final manuscript for important intellectual content. All authors approved the final manuscript.

## Patient and public involvement

Patients or members of the public were not involved in the design, conduct, analysis, interpretation, or reporting of this study

## Patient consent for publication

Not applicable

## Ethics approval

Ethical approval was obtained from the Ethics Review Committee, Faculty of Medicine and Allied Sciences, Rajarata University of Sri Lanka (ERC/2024/60).

## Data availability statement

Data are available on reasonable request. Please contact Dr. Manori Vijaya Kumari.

